# Systemic immune profile in hospitalized patients with tuberculosis and Covid-19

**DOI:** 10.1101/2025.06.05.25329097

**Authors:** Felipe M Ridolfi, Sabrina Soares Guimarães, Mariana Araujo-Pereira, Naíse S Bemfica, Quézia M de Oliveira, Mariana S Xavier, Eric H Roma, Flavia M Sant’Anna, Alice MS de Andrade, Marina C Figueiredo, Timothy R Sterling, Bhavna G Gordhan, Bavesh D Kana, Bruno B Andrade, Adriano Gomes-Silva, Valeria C Rolla, the Associative BRICS Research in Covid-19 and TB (ABRICOT) Research Group

**Author notes:** Corresponding author: **Valeria Cavalcanti Rolla, M.D., Ph.D.** **|**, Alternative corresponding author: **Felipe Moreira Ridolfi, M.D., Ph.D.** **|**. These authors contributed equally to this work and share first authorship. These authors contributed equally to this work and share senior authorship. **Associative BRICS Research in Covid-19 and TB (ABRICOT) Research Group** (alphabetical order): Alessandra Saboia, Aline Benjamin, André Miguel Japiassu, Andrea Gina Varon, Carolina Arana Stanis Schmaltz, Eduardo Martins Gama, Francine Peixoto Ignácio Cavalcanti, Gabriela Corrêa e Castro, Georgia Claudia Trade Santos, João Victor do Nascimento, Juliana do Carmo Godinho, Marcella de Oliveira Belém, Leonardo Teodosio de Mello, Maria Cristina Lourenço, Maria Luciana Silva de Freitas, Mirella Gomes Balbino, Monique Pinto Gonçalves, Paulo Leandro Garcia Meirelles Junior, Ramaiana Hackbart Timm, Renata Marialva, Renne Kós Rocque, Rosa Placido Pereira, Samyra Almeida da Silveira, Thais Teixeira Machado, and Thiago da Silva Paulo.

## Abstract

Tuberculosis (TB) and Covid-19 are respiratory diseases and their interaction could increase lung impairment and mortality. However, few cases of concomitant TB and Covid-19 have been reported, and biomarkers are still poorly described. This cross-sectional study included in-hospital adult (>18 years old) participants, with pulmonary TB (TB group), with Covid-19 (Covid-19 group), and with TB/Covid-19 co-infection (TB/Covid-19 group). We compared baseline demographic, clinical, laboratory, and inflammatory profile features across the groups. Inflammatory soluble factors were assessed via MILLIPLEX MAP Human Cytokine/Chemokine Premixed 29 Plex and Luminex Intelliflex xMAP. We used the Chi-square test for categorical variables, and the Mann-Whitney *U* or the Kruskal-Wallis tests for continuous variables. A random forest model evaluated the variables importance in discriminating the groups, assessed by the Mean Decrease Gini Index (≥1.5). Among the 76 participants included, 33 (43%) with pulmonary TB, 13 (17%) with TB/Covid-19, and 30 (39%) with Covid-19. Male participants were predominant in all groups. The TB and TB/Covid-19 groups had lower median body mass index (BMI) (18.4 [interquartile range (IQR) 16–22] and 19.4 [IQR 16.1–20.3], respectively) compared to the Covid-19 group (25.5 [IQR 22.0–31.7]; p<0.001). People living with HIV/AIDS were more frequent in TB (63%) and TB/Covid-19 (61%) groups than in Covid-19 (20%; p=0.001). Participants in the Covid-19 group had significantly more comorbidities (63%) than those in the TB (9%) and TB/Covid-19 (15%) groups (p<0.001). Comorbidities included HIV seropositivity, diabetes mellitus, chronic obstructive pulmonary disease, and hypertension. Cavitation on chest CT scan was observed in 45% of TB and 61% of TB/Covid-19 participants, but none in the Covid-19 group (p<0.001). Covid-19 participants exhibited lower platelet counts (p=0.004), higher creatinine (p<0.001), and higher urea levels (p=0.004). Random forest analysis identified urea, hemoglobin, hematocrit, and platelets as the best tests to discriminate TB/Covid-19 from the other groups (Area Under the Curve>0.70 in both cases). MCP-1 was significantly elevated in Covid-19 compared to TB and TB/Covid-19 (p=0.014). No inflammatory signature differentiated TB/Covid-19 from single infections. TB and Covid-19 are clinically similar and TB/Covid-19 co-infection is uncommon. Overall, the plasma inflammatory response could not differentiate Covid-19, TB and TB/Covid-19. Comorbidities, radiologic findings, and laboratory tests, such as urea, hemoglobin, hematocrit and platelets, may be useful tools to distinguish TB and TB/Covid-19, as well as Covid-19 and TB/Covid-19.

## 1. Introduction

The Covid-19 pandemic negatively impacted health services, including the diagnosis and notification of other infectious diseases, such as tuberculosis (TB). (1–4) Although Brazil experienced many waves of Covid-19 with different SARS-CoV-2 strains since January 2020, widespread vaccination decreased the incidence and the severity of Covid-19 cases. (5) TB, like Covid-19, is an airborne pathogen that most commonly affects the lungs. Clinically, TB and Covid-19 may be indistinguishable, especially in mild cases as both diseases present with similar respiratory symptoms (6)

Several human inflammatory markers have been studied as predictors of Covid-19 disease severity. These include C-reactive protein (CRP), D-dimer, ferritin, interleukin (IL)-6, troponin, lymphocyte count, absolute number of CD4+ and CD8+ T lymphocytes, and neutrophil-lymphocyte ratio (NLR). (7–10) In severe cases, consistent increase in CRP, D-dimer, erythrocyte sedimentation rate (ESR), lactic dehydrogenase (LDH), alanine aminotransferase (ALT), aspartate aminotransferase (AST), and a reduction in absolute lymphocyte count has been observed. (11) TB elicits some common inflammatory markers with Covid-19 – i.e., elevated CRP, neutrophils, lymphocytes, ESR, and IL-6, but can also present with elevated levels of interferon-gamma (IFN-γ) and platelet count. (12)

Since co-infection with *Mycobacterium tuberculosis* (Mtb) and SARS-CoV-2 is complex and still unclear, in this study, we aimed to understand the clinical and laboratory characteristics of TB/Covid-19 coinfection compared to the respective individual diseases. We assessed clinical and biomarker differences, aiming to better discriminate TB, Covid-19 and TB/Covid-19.

## 2. Material and methods Study Design and Setting

The cross-sectional Associative BRICS Research in Covid-19 and TB (ABRICOT) study was set up to investigate the effect of Covid-19 on TB immunopathogenesis. The Laboratório de Pesquisa Clínica em Micobacterioses (LAPCLIN-TB) recruited in-hospital participants in the Covid-19 Hospital Center, at the Instituto Nacional de Infectologia Evandro Chagas, Fiocruz, Rio de Janeiro, Brazil, between November 26^th^, 2020, and May 10^th^, 2023. This large tertiary and referral center, with 190 beds at that time, was built in three months during the Covid-19 pandemic to assist Covid-19 cases (and other infectious disease) from all over the Rio de Janeiro state, Brazil.

### Ethics Statement

The ABRICOT study was approved by the institutional review board of the Instituto Nacional de Infectologia Evandro Chagas (CAAE: 38791120.8.0000.5262). Written informed consent was obtained from all participants, and all clinical investigations were conducted according to the principles expressed in the Declaration of Helsinki.

### Study Population

We included adult participants (>18 years old), with pulmonary TB (TB group), or Covid-19 (Covid-19 group), and participants with concomitant TB and Covid-19 (TB/Covid-19 group). We excluded participants receiving TB treatment for >1 week before enrollment, pregnant or lactating participants, and drug-resistant (DR)-TB cases.

Covid-19 was diagnosed by a positive SARS-CoV-2 real-time polymerase chain reaction test from a nasopharyngeal swab or with a positive nasopharyngeal SARS-CoV-2 immunochromatographic antigen test. Pulmonary TB was diagnosed by a positive Xpert-MTB-RIF Ultra test and/or culture (growing Mtb) from respiratory samples. TB/Covid-19 cases had both diseases diagnosed within a 14-day interval.

### Variables and definitions

We considered baseline demographic and clinical variables for all groups, such as age and sex, cough, cough duration, fever, fatigue, night sweats, weight loss, chest pain, headache, dyspnea, anorexia, purulent sputum, hemoptysis, odynophagia, dysphagia, and hoarseness. We accounted for the following comorbidities: HIV (based on serology); diabetes mellitus, based on glycated hemoglobin (HbA1c) >6.5% – participants with HbA1c between 5.7-6.4% were defined as having pre-diabetes. Chronic obstructive pulmonary disease (COPD)/emphysema, bronchitis, hypertension (self-reported), hepatitis B virus (HBV) (based on serology), and obesity were grouped as other comorbidities. Despite the comorbidities, none of the participants were on immunosuppressive therapy at the time of recruitment.

We considered laboratory parameters such as hematocrit, hemoglobin, white blood cell count, ALT, AST, gamma glutamyl transferase (GGT), alkaline phosphatase (ALP), and bilirubin, kidney function (creatine, urea), and inflammatory markers, such as CRP, LDH, and ESR.

Variables selected to explore TB characteristics were body mass index (BMI), disseminated TB (defined by concomitant pulmonary and extrapulmonary disease), presence of pulmonary cavitation on chest computed tomography scan (CT). For Covid-19 participants (in both the TB/Covid-19 and Covid-19 groups) the laboratory parameters were CRP, ferritin, and LDH. (16)

Covid-19 vaccination in Brazil started in 2021, initially with CoronaVac and AstraZeneca vaccines. Limited quantities of these vaccines resulted in many people in 2021-2022 receiving booster vaccines that were different from the initial vaccine. (13) For this study, a Covid-19 vaccination cycle was considered complete if the individual received at least three doses of any of the available vaccines in Brazil.

### Luminex assay

Plasma was derived from blood samples obtained at the baseline visit and stored at −80°C. These samples were used for quantification of inflammatory soluble factors by the MILLIPLEX MAP Human Cytokine/Chemokine—Premixed 29 Plex using Luminex Intelliflex xMAP technology to define the systemic inflammatory response for each participant. This allowed for the simultaneous quantification of the following cytokines, chemokines or growth factors: EGF, Eotaxin, G-CSF, GM-CSF, IFN-α2, IFN-γ, IL-10, IL-12(p40), IL-12(p70), IL-13, IL-15, IL-17, IL-1RA, IL-1α, IL-1β, IL-2, IL-3, IL-4, IL-5, IL-6, IL-7, IL-8, IP-10, MCP-1, MIP-1α, MIP-1β, TNF-α, TNF-β, and VEGF. Measurements were taken following the manufacturer’s instructions.

### Statistical Analysis

Descriptive statistics were used to present the data. Continuous variables were presented as median values with 25% and 75% interquartile ranges (IQR), whereas categorical variables were described using frequency and proportions (%). The Chi-square test was used to compare categorical variables between study groups, and the Mann-Whitney U test (for two unmatched groups) and the Kruskal-Wallis test (for three groups) were used to compare continuous variables.

The inflammatory soluble factor data were log-transformed to evaluate the overall inflammatory response, and an unsupervised hierarchical cluster analysis (Ward’s method) with dendrograms representing the Euclidean distances was performed.

A random forest model was applied to evaluate the importance of biochemical and immunological parameters in distinguishing the different participant groups. Variables included cytokines and chemokines. The model was trained using standard procedures for classification, and variable importance was assessed based on the Mean Decrease in Gini Index, a measure of how much each variable contributed to the model’s accuracy. The threshold for variable selection was set at a Mean Decrease Gini value of ≥ 1.5, ensuring that only the most relevant discriminants were considered.

To assess the discriminative power of markers with an MDI≥1.5, Receiver Operating Characteristic (ROC) curves were generated. The area under the curve (AUC) was calculated for each pairwise classification model to evaluate sensitivity and specificity. Higher AUC values indicated better classification performance, and confidence intervals for AUC values were computed to assess the robustness of the models. All statistical analyses were performed using R software (version 4·4·1), with a 0.05 significance level.

## 3. Results

A total of 92 hospitalized participants were enrolled, with 76 (82.6%) included in the analysis. This included 33 in the TB group, 13 in the TB/Covid-19 group, and 30 in the Covid-19 group. Four participants from the Covid-19 group were excluded as they had suspected TB during clinical evaluation, 6 months after inclusion into the study. Twelve participants with suspected TB were included temporarily in the TB group but were subsequently excluded due to negative results of both the Xpert MTB/RIF Ultra and Mtb culture. (**Fig. 1**).

**Fig 1.**
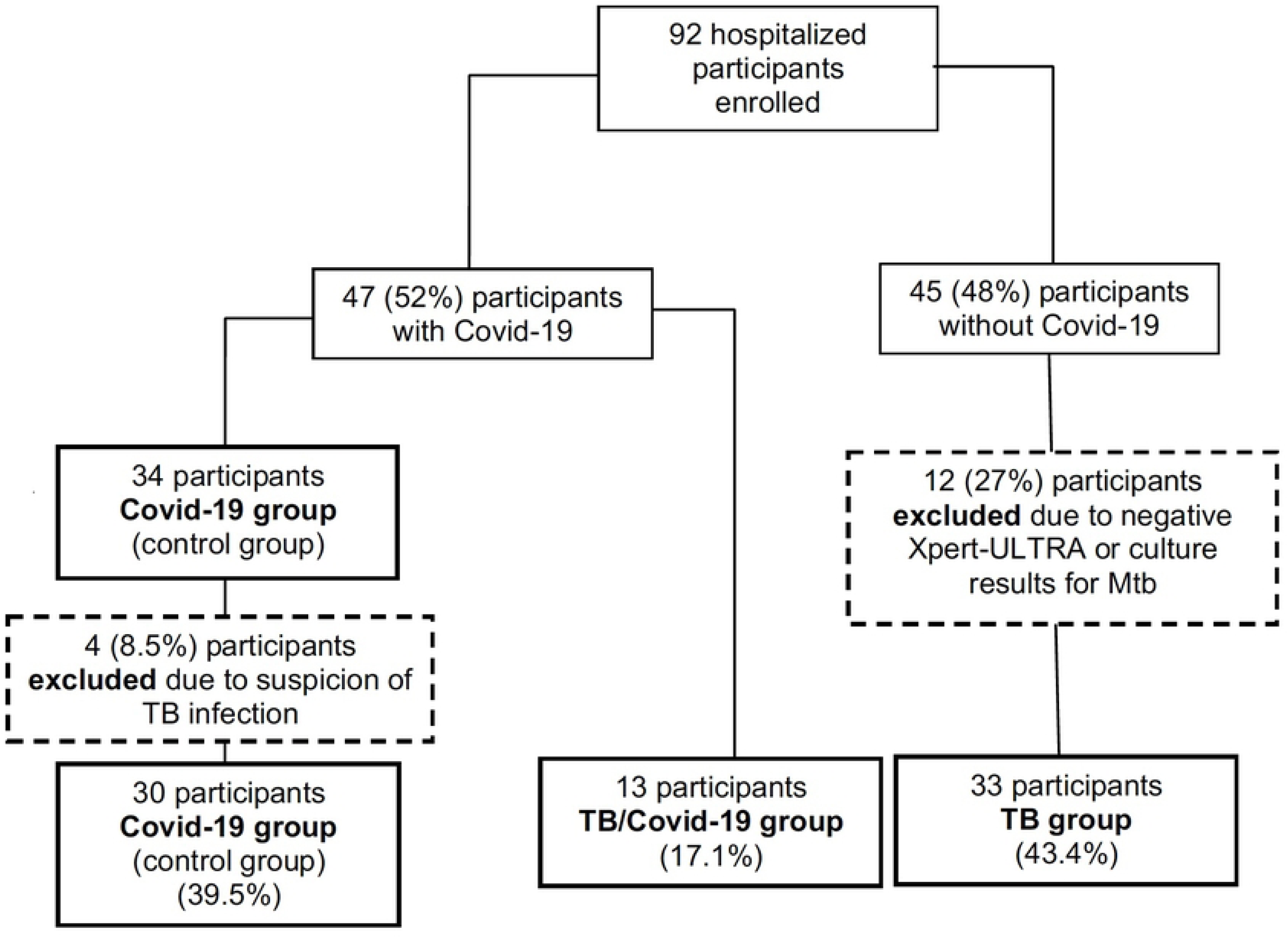
Participant disposition flow chart for individuals recruited in this study and the reasons that led to exclusion of participants.

The median age was 37 years (interquartile range [IQR] 30–48] for the TB group, 41 years (IQR 26–55) for TB/Covid-19, and 59 years (IQR 45.5–70.8) for Covid-19 (p=0.267). Male participants were predominant in all groups. TB and TB/Covid-19 groups had lower BMI (18.4 [IQR 16-22] and 19.4 [IQR 16.1-20.3], respectively) compared to Covid-19 (25.5 [IQR 22.0-31.7]) (p<0.001). People living with HIV/AIDS (PLWHA) were more frequent in the TB (64.6%) and TB/Covid-19 (61.5%) groups than the Covid-19 (20%) group (p=0.001). Most (63.3%) Covid-19 participants had comorbidities, unlike the TB (9%) and TB/Covid-19 (15.4%) groups (p<0.001). Cavitation on chest CT was observed in TB (45%) and TB/Covid-19 (61.5%) groups but not in Covid-19 (p<0.001). Covid-19 vaccination was low across all groups, with TB and TB/Covid-19 participants more likely to be unvaccinated or incompletely vaccinated (p=0.011) (**Table 1**).

**Table 1.**
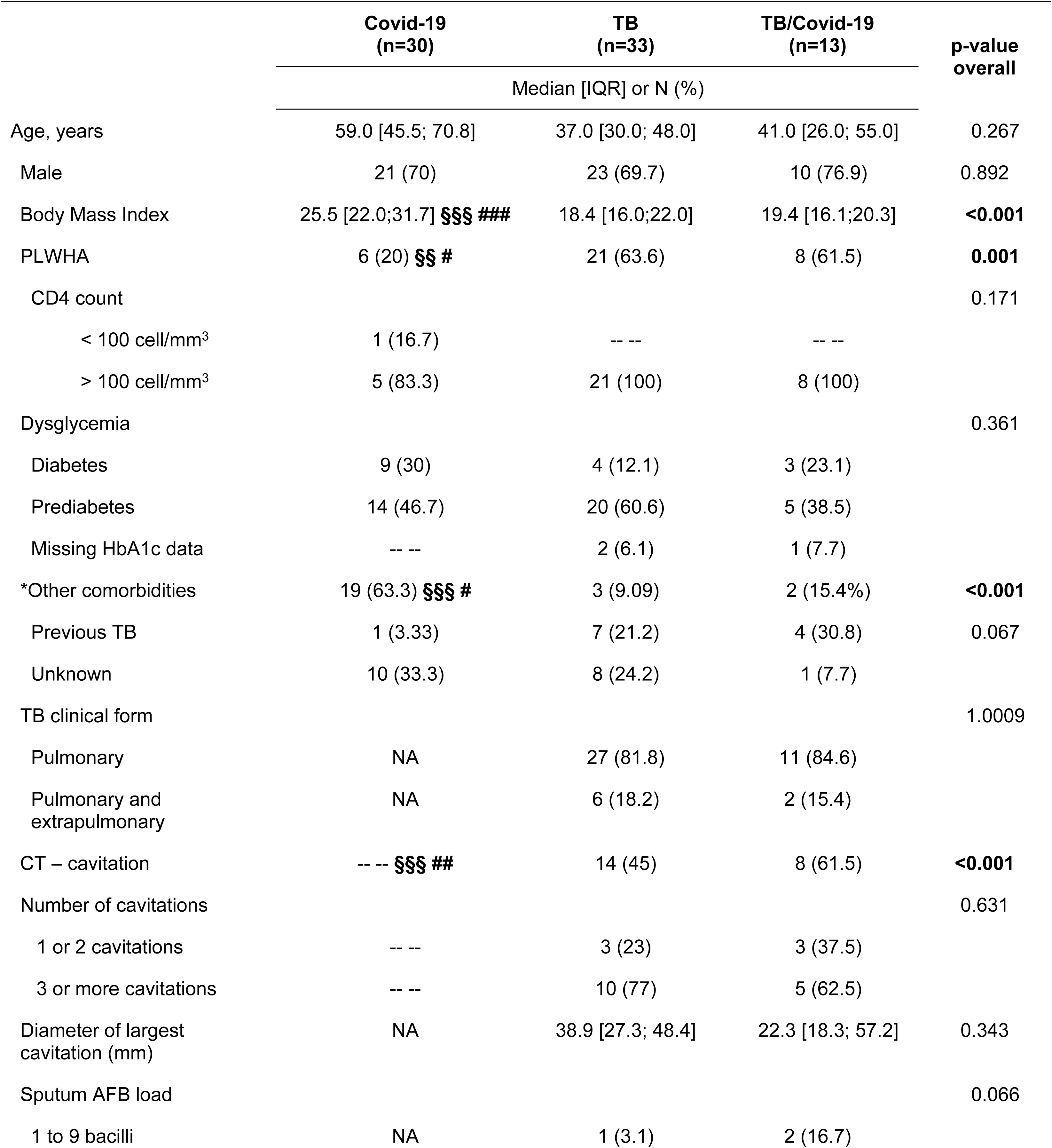
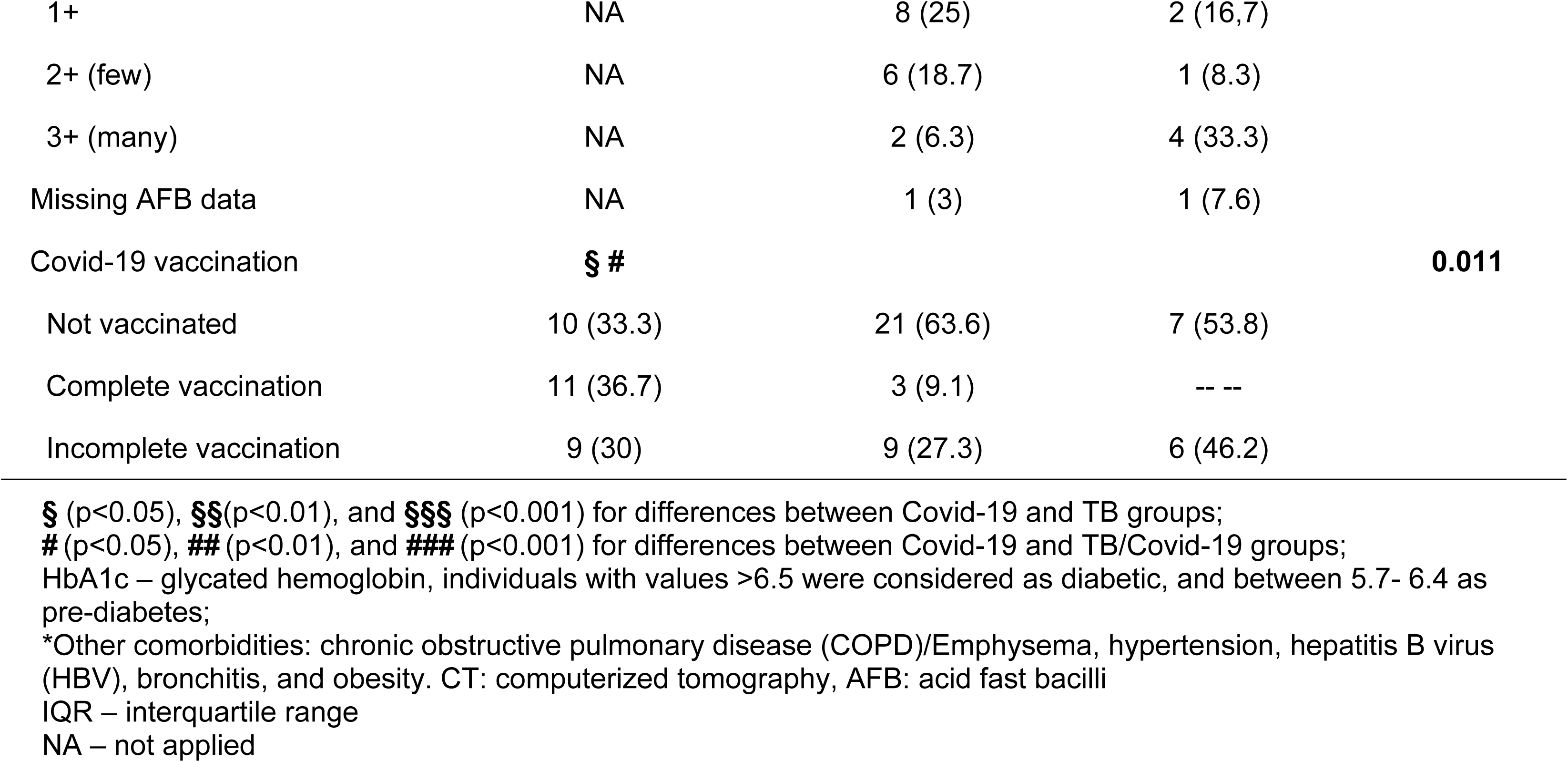
Demographic and clinical characteristics of Covid-19, tuberculosis (TB) and TB/Covid-19 coinfection groups at the baseline visit.

In comparison to Covid-19 participants, TB and TB/Covid-19 patients had more weight loss (p<0.001 for both), longer cough duration (p<0.001 and p<0.01 respectively), more anorexia (p<0.01 for both) and more purulent sputum (p<0.05 for both). TB participants had more fatigue (p<0.01) but less fever (p<0.05) and less odynophagia (p<0.05) than Covid-19 participants (**Table 2**).

**Table 2.** Signs and symptoms registered at the baseline visit of hospitalized Covid-19, tuberculosis (TB), and TB/Covid-19 coinfection at baseline visit.

| Signs and Symptoms | Covid-19<br>(n=30) | TB<br>(n=33) | TB/Covid-19<br>(n=13) | p-value<br>overall |
| --- | --- | --- | --- | --- |
|  | Median [IQR] or N (%) |  |  |  |
| Cough | 21 (69) | 25 (76.1) | 9 (68.2) | 0.805 |
| Cough duration (weeks) | 1.0 [1.0; 2.0] **§§§ ##** | 4.0 [2.0; 8.0] | 4.0 [2.5; 9.0] | **<0.001** |
| Fever | 14 (45.7) **§** | 25 (77.1) | 12 (69.2) | **0.047** |
| Weight loss | 8 (25.7) **§§§ ###** | 28 (86.5) | 12 (92.3) | **<0.001** |
| Fatigue | 20 (65.7) **§§** | 29 (89.6) | 9 (68.2) | **0.003** |
| Chest pain | 7 (22.3) | 7 (19.9) | 2 (13.4) | 1.000 |
| Night sweats | 3 (9) | 7 (19.9) | 2 (13.4) | 0.433 |
| Headache | 5 (15.7) | 1 (18.2) | 1 (5.7) | 0.183 |
| Adenomegaly | -- -- | 4 (11.5) | 2 (14.4) | 0.056 |
| Anorexia | 7 (22.3) **§§ ##** | 21 (64.6) | 10 (75.9) | **<0.001** |
| Dyspnea | 20 (65.7) | 24 (74) | 5 (37.5) | 0.119 |
| Purulent sputum | 5 (15.7) **§ #** | 13 (38.6) | 6 (45.2) | **0.046** |
| Hemoptysis | 1 (2.3) | 2 (4.3) | 1 (6.7) | 0.816 |
| Odynophagia | 5 (15.7) **§** | -- -- | 2 (14.4) | **0.033** |
| Runny nose | 4 (13.8) | -- -- | -- -- | 0.112 |
| Loss of smell and or taste | 3 (10.3) | -- -- | 1 (4.7%) | 0.249 |

**§** (p<0.05), **§§**(p<0.01), and **§§§** (p<0.001) for differences between Covid-19 and TB groups; **#** (p<0.05), **##** (p<0.01), and **###** (p<0.001) for differences between Covid-19 and TB/Covid-19 groups;

TB and TB/Covid-19 groups presented with lower hematocrit and hemoglobin levels compared to the Covid-19 group (p=0.017 and 0.008, respectively). The white blood cell count was similar among all three groups, with no significant differences in the differential count. In relation to Covid-19 participants, TB participants were more likely to have altered liver function, mainly through ALP and conjugated bilirubin elevation (p<0.05 for both). However, the Covid-19 group had lower platelet count (p=0.004), and higher levels of creatinine (p<0.001) and urea (p=0.004), compared to the TB and TB-Covid-19 groups. The inflammatory marker CRP was above the reference value in all groups, but the TB and TB/Covid-19 groups had at least a 2-fold higher plasma CRP than the Covid-19 group (**Table 3**).

**Table 3.**
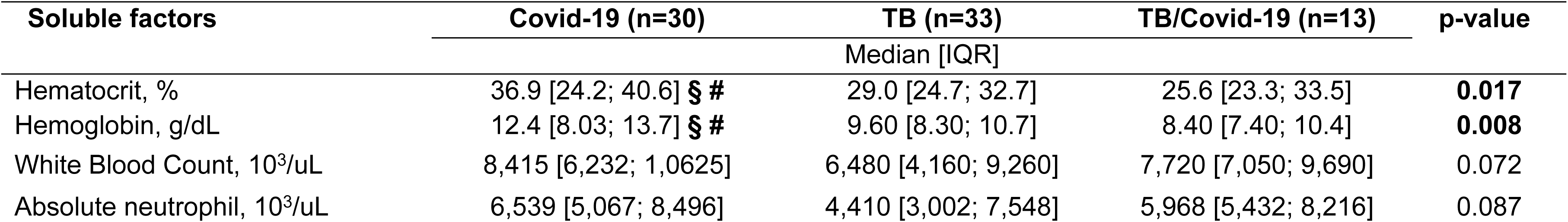
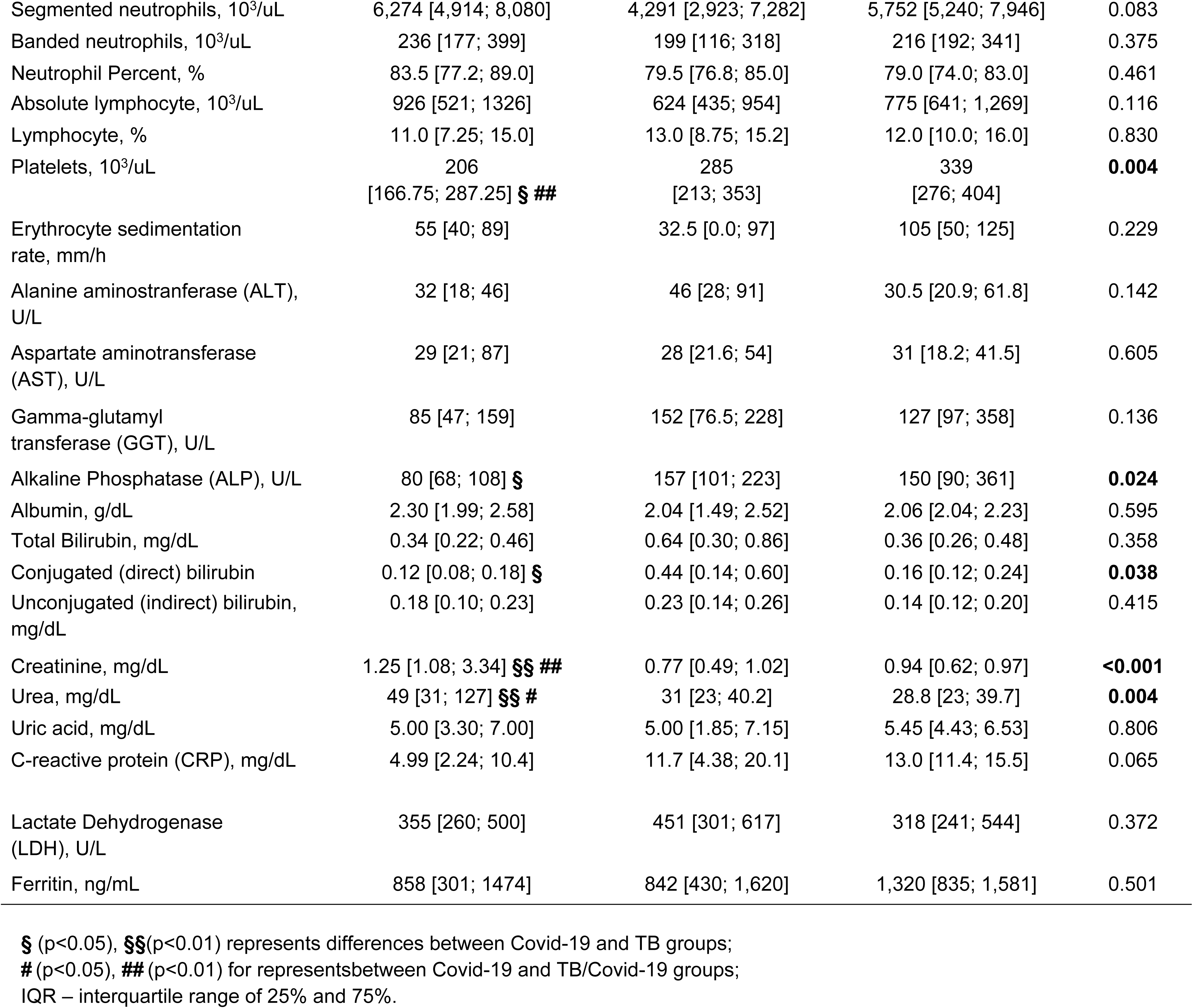
Hematologic and biochemistry parameters in Covid-19, tuberculosis (TB) and TB/Covid-19 groups at the baseline visit.

The random forest model identified biochemical parameters as the most important determinants in distinguishing patient groups. Among the variables, urea, hemoglobin, hematocrit and platelets showed the highest Mean Decrease in Gini Index, with values above 1.5. In contrast, most of the inflammatory soluble factors, such as IL-7, IL-10, and TNFα, had lower importance, with Mean Decrease Gini values below 0.5 (**Fig 2A**). **Fig 2B** presents Receiver Operating Characteristic (ROC) curves comparing the ability of urea, hemoglobin, hematocrit and platelet combined to distinguish between the groups. The Area Under the Curve (AUC), along with sensitivity and specificity values, is reported for each classification model, reflecting the discriminatory power of this marker-based signature (**Supplementary Table 2**). This signature is particularly useful to distinguish between TB and TB/Covid-19 (AUC: 0.72 95%CI 0.53-0.90, p = 0.023); as well Covid-19 and TB/Covid-19 (AUC: 0.84 95%CI 0.69-0.99, p<0.001).

**Fig 2.**
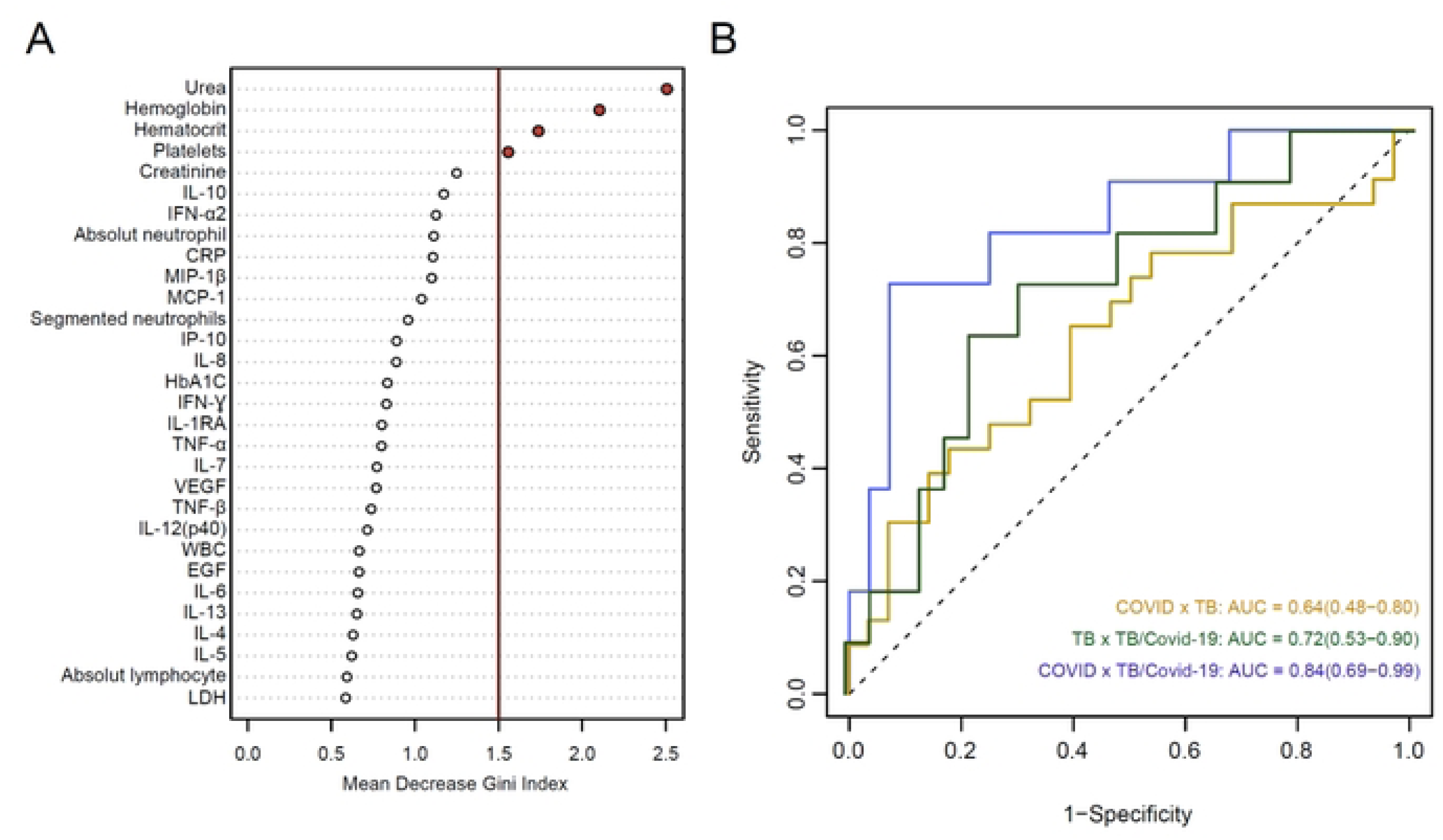
(A) Random forest analysis of variable importance distinguishing TB, Covid-19, and TB/Covid-19 groups. Fig 2. (B) Receiving Operation Curves illustrate the discriminatory power of selected markers (urea, hemoglobin, hematocrit, and platelets) for differentiating groups.

This figure demonstrates the importance of the different variables based on the Mean Decrease in Gini Index, indicating their relative contributions to distinguish groups. Urea, hemoglobin, hematocrit, and platelets are highlighted as the most important variables with an Index greater than 1.5. The Area Under the Curve (AUC) are displayed for each classification model. The AUC metrics are described in Supplementary Table 2. The only significant difference was Covid-19 participants having higher levels of MCP-1 in relation to the TB and TB/Covid-19 groups (p=0.014) (**Supplementary Table 1**). Additionally, the heatmap analysis did not show an inflammatory signature based on soluble factors that could differentiate the TB/Covid-19 group from the TB or Covid-19 groups. Furthermore, within each group, it was not possible to associate the observed clustering with any other patients’ conditions, especially in relation to PLWHA (**Fig 3**).

**Fig 3.**
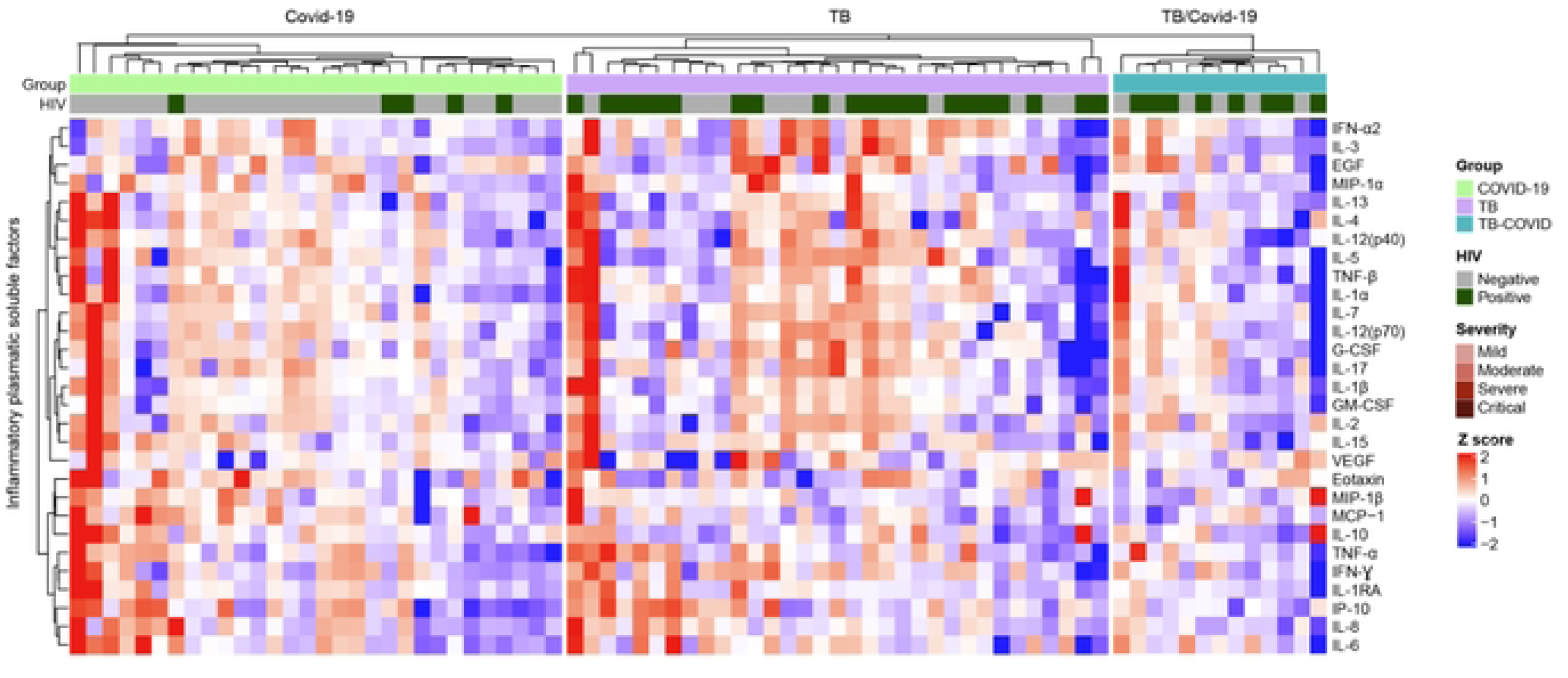
Hierarchical clustering analysis of soluble factors in plasma samples from TB, COVID-19, and TB/COVID-19 participants by HIV status.

Analysis of the soluble factors quantified by the Luminex assay in plasma samples obtained at the baseline visit from participants in the Covid-19 (light green horizontal bar), TB (purple horizontal bar) and TB/Covid-19 (blue horizontal bar) groups. The second horizontal bar identifies participants living with HIV/AIDS or not. The third horizontal bar identifies participants with mild, moderate, severe or critical disease. Heat map with a hierarchical cluster analysis can be seen at the top of the figure (Ward’s method, with dendrograms implying the Euclidean distance) The concentration of soluble factors can be visually determined based on the intensity of the colors deep red (highest concentration) and deep blue (lowest concentration). Each column represents one participant.

## 4. Discussion

In this cross-sectional study, we evaluated the demographic and clinical, radiological, and laboratory parameters, together with plasma inflammatory response, among TB, TB/Covid-19, and Covid-19 participants. Despite similar clinical presentations, and no significant differences in immunological markers, these participants had some findings related to hematological and biochemical parameters that may be used to differentiate the groups.

Symptoms such as cough, headache, and dyspnea were similar between all groups. However, compared to participants with Covid-19, individuals with TB and TB/Covid-19 had a longer duration of cough, weight loss, anorexia, and purulent sputum. These signs and symptoms are commonly seen in patients with TB, whereas patients with Covid-19 usually have acute disease, with dry cough and dyspnea. (14)

At the start of the Covid-19 pandemic, it was expected that TB/Covid-19 would be a highly frequent disease association. However, in our study cohort TB/Covid-19 only thirteen participants (14.1%) had both diseases. Several studies from 2019 and 2020 evaluating the prevalence of co-infections among hospitalized patients with Covid-19 reported no cases of TB/Covid-19 coinfection. (15) In a subsequent cohort study, from eight countries, assessment of TB and Covid-19 in 49 patients showed that TB was diagnosed before Covid-19 in 26 patients (53%), Covid-19 was diagnosed before TB in 14 individuals (28.5%) while the concomitant diagnosis within the same week was found in only 9 patients (18.3%). This finding demonstrates that TB/Covid-19 co-infection was not as frequent as initially hypothesized. (16)

Compared to TB and TB/Covid-19, Covid-19 participants had a higher BMI, had more comorbidities, such as hypertension, and COPD, and were less likely to be PLWHA. These findings are consistent with a study that evaluated 5,700 hospitalized patients with Covid-19 in the New York City area, in whom the most common comorbidities were hypertension, obesity, and diabetes (17), whilst in another study, critically ill patients with Covid-19 showed a high prevalence of COPD, chronic kidney disease, and congestive heart failure. (18) This finding is also supported by a Chinese study, which found a 48% comorbidity prevalence among Covid-19 participants, with hypertension being the most common, followed by diabetes, and coronary heart disease. (19)

PLWHA were more prevalent in TB and TB/Covid-19 groups than the Covid-19 group, showing an association with TB and not with Covid-19. (20,21) This finding is supported by a large study with more than 200,000 patients, which showed 9.1% of PLWHA among hospital-admitted Covid-19 patients. (21) Initially, researchers expected to see more PLWHA among Covid-19 cases due the immunodeficiency caused by HIV, but HIV-infection was infrequent among Covid-19 cases. (17–19,21) However, the same studies showed that HIV-infection has been associated with higher mortality among Covid-19 patients with CD4<200 cells/μL, with HIV viral load of 1,000 copies/μL or more. (21)

As a viral disease, Covid-19 is expected to present with lymphopenia and thrombocytopenia. Although the white blood cell count did not differ significantly among the groups, the Covid-19 group had lower platelet counts, compared to TB and TB/Covid-19, which is as expected. (17,22–24) In the Mean Decrease Gini Index, urea, hemoglobin, hematocrit and platelets were the variables that most discriminated the groups. The AUC was able to distinguish between TB and TB/Covid-19, along with Covid-19 and TB/Covid-19 (AUC: 0.84 95%CI 0.69-0.99, p<0.001). Covid-19 participants had higher levels of creatinine, some of them with renal function impairment, compared to TB and TB/Covid-19 participants. As a multi-system disease, Covid-19 has been associated with impairment of renal function, especially among in-hospital patients. (17,19)

In our study, the Covid-19 group did not present with cavitation on chest CT scan, which is in accordance with the literature, in which the common radiographic alterations of Covid-19 are reticulonodular and ground-glass opacities, rather than cavitation. (18,19) Surprisingly, the levels of CRP, LDH, ferritin, and the plasma inflammatory response were similar between the groups, except for higher MCP-1 levels among Covid-19 participants. This marker has been associated with disease severity and as a risk factor for death in Covid-19 patients. (25) It is important to emphasize that the systemic immune response assessed by plasma levels of cytokines, chemokines and growth factors can be influenced by other stimuli associated with comorbidities and other infections, although in our study this was not observed. (26,27) A previously published study showed that a TB/Covid-19 diagnosis (without other infections such as HIV) was statistically associated with higher levels of TNF-α. (28) Similarly, we expected to identify a potential biomarker that would differentiate TB/Covid-19 from TB cases; and although MCP-1 was statistically significant, it could not to discriminate TB-Covid from TB. Overall, the vaccination was low. The vaccination was more frequent in the Covid-19 group. The study started in 2020, when the vaccination was initiated in Brazil by age groups. Elderly persons (>60 years-old) and healthcare professionals were prioritized and young adults started vaccination later. Most participants included in our study were young and were vaccinated later in 2021.

There were some limitations of our study. This includes the small sample size, particularly for the TB/Covid-19 group, that could have impacted on the findings. The D-dimer test was performed for all participants, however, due to different laboratory techniques and different cutoff points, we were unable to include this variable in the analysis. On the other hand, we highlight the study recruited hospitalized participants in a large tertiary referral center for Covid-19 management in the state of Rio de Janeiro during the pandemic, and the extensive inflammatory profile we performed were strengths of our study.

Based on TB and Covid-19 being clinically similar, we hypothesized that TB/Covid-19 coinfection would be common. However, in our cohort this association was uncommon. Whilst the systemic inflammatory response could not differentiate TB and TB/Covid-19, urea, hemoglobin, hematocrit and platelets may be useful tools to distinguish TB and TB/Covid-19, as well as Covid-19 and TB/Covid-19, supporting the hypothesis that biochemical parameters are more robust in discriminating the groups than immunological markers.

## Data Availability

The datasets used and analyzed for the current study will be deposited at the Fiocruz Institutional Repository, ARCA, and a link will be available after acceptance for publication.

## 6. Author Contributions

SSG - recruited patients, prepared data for analysis, and revised the final version of manuscript; MAP - curated the Luminex output data, assisted with data analysis and design of figures and revised the manuscript; QMO - supervised recruitment, managed data quality and ethical regulatory documents; MSX - revised the final version of the manuscript; EHR – participated in the discussion of results and interpretation of data; NB - recruited participants and collected the biological samples; FMSA - recruited participants and discuss the results; FMR – recruited participants, discuss the results and wrote the manuscript; AMSA - performed the Luminex experiments and revised the final version of the manuscript; MCF - worked on the protocol, and revised the manuscript; TRS - interpreted the data and revised the manuscript; BGG and BDK - wrote the main protocol and revised the manuscript; BBA - supervised data analysis, figures design, and revised and provided inputs the manuscript; AGS - supervised sample storage, curated the samples for Luminex analysis, wrote the protocol, data interpretation and wrote the manuscript; VCR - wrote the protocol, supervised recruitment, established the initial concept and wrote the manuscript.

## Acknowledgments

The authors thank all the study participants and the Associative BRICS Research in Covid-19 and TB (ABRICOT) Group.

